# Subtle white matter intensity changes on FLAIR imaging in patients with ischemic stroke

**DOI:** 10.1101/2023.05.17.23290149

**Authors:** Pedro Cougo, Heber Colares, João Gabriel Farinhas, Mariana Hämmerle, Pedro Neves, Raquel Bezerra, Alex Balduíno, Ona Wu, Octavio M. Pontes-Neto

## Abstract

**Background:** There is increasing evidence from pathology and neuroimaging suggesting that the structural abnormalities which characterize leukoaraiosis are present within regions of normal-appearing white matter. In this study, we aimed to verify whether signal intensity on FLAIR imaging in normal-appearing white matter is related to the leukoaraiosis burden.

**Methods:** We performed a cross-sectional study of adult patients admitted with a diagnosis of acute ischemic stroke or transient ischemic attack. Leukoaraiosis was segmented using a semi-automated method involving manual outlining and signal thresholding. White matter regions were segmented based on the probabilistic tissue maps from the International Consortium for Brain Mapping 152 atlas. Also, white matter was further segmented based on voxel-distance from leukoaraiosis borders. Normalized mean FLAIR signal intensity on normal-appearing white matter (NAWM_M_) was used as a dependent variable in univariate and multivariate statistical analysis, and leukoaraiosis volume quartiles (LKA_V_) and clinical data as independent variables.

**Results:** One-hundred consecutive patients were selected for analysis (53% female, mean age 68 years). NAWM_M_ was higher in the vicinity of leukoaraiosis and progressively lower at increasing distances from leukoaraiosis. NAWM_M_ was independently associated with leukoaraiosis volume. In voxels in the vicinity of leukoaraiosis borders, there was a linear association between LKA_V_ and NAWM_M_ (*B*=0.03; *P*<0.01). In voxels non-adjacent to leukoaraiosis borders, there was a non-linear association, with higher values of NAWM_M_ among patients in the second and third quartiles, and lower in the first and fourth quartiles (*B*=-0.21 for the second order term; *P*<0.01). In multivariate analysis, leukoaraiosis was the only variable independently related to NAWM_M_.

**Conclusions:** Our results show that normal-appearing white matter exhibits subtle signal intensity changes that are related to leukoaraiosis burden. The neuroimaging signature of these subtle changes suggests that they might not be a milder form of leukoaraiosis, but rather rather reflect widespread etiopathogenic processes underlying small-vessel disease and thus, leukoaraiosis development.

## Introduction

Leukoaraiosis is a neuroimaging marker of small-vessel disease that is highly prevalent and associated with stroke incidence, cognitive decline and increased mortality.^1,2^ Leukoaraiosis has been associated with well-established vascular risk factors, such as age and hypertension, as well as clinical and tissue outcomes after acute stroke, including penumbral tissue loss, long-term functional outcome and stroke recurrence.^3–9^

Leukoaraiosis is defined as conspicuous regions of brain white matter that are hyperintense on T2-weighted MRI and that typically follow either a periventricular or a subcortical, patchy distribution.^10^ Leukoaraiosis is especially conspicuous on the fluid-attenuated inversion recovery (FLAIR) MRI sequence. However, it has been shown that leukoaraiosis-related changes in both pathology and neuroimaging can also be found in normal-appearing white matter on T2-weighted images. Studies using diffusion tensor imaging (DTI) techniques found abnormal diffusion values in normal-appearing white matter in patients with more severe leukoaraiosis.^11,12^ In studies evaluating blood-brain barrier disruption with dynamic contrast-enhanced and perfusion MRI, patients with small-vessel disease presented with abnormalities in normal white matter that were also related to the burden of leukoaraiosis.^13,14^ Moreover, both diffusion changes and blood-brain barrier disruption in normal-appearing white matter seem to be related to clinical outcomes after an acute ischemic stroke.^15,16^

It appears therefore that the underlying neuroimaging changes related to small-vessel disease are not only represented by the more conspicuous hyperintensity of leukoaraiosis but also by subtle, diffuse and clinically relevant changes on what appears to be normal white matter on T2-weighted MRI. In this study, we aimed to verify whether the signal intensity on FLAIR imaging in normal-appearing white matter is related to the burden of leukoaraiosis in patients with ischemic stroke.

## Material and methods

### Subjects

This study was approved by our Institutional Review Board with a waiver of informed consent. We retrospectively selected eligible patients from a prospective, hospital-based, clinical stroke registry. The registry screens all adult patients admitted to our institution with a clinical diagnosis of stroke of any type. For this study, we reviewed patients from January, 2020 to July, 2022, who were admitted with a diagnosis of ischemic stroke or transient ischemic attack, and who had an MRI performed at any time during the hospital admission, including FLAIR and diffusion-weighted imaging (DWI) imaging, and either T2- or T1-weighted imaging. Patients with bilateral infarcts (either acute or chronic) or motion artifacts were excluded.

### Imaging acquisition and analysis

MRI was performed on a 3T Siemens, Skyra scanner. Echo time, repetition time and inversion time for FLAIR images were 72, 9000 and 2500 ms. Slice thickness and field-of-view were 3.5 mm and 21 × 24 cm or 18 × 24 cm. Imaging analysis was performed with the Medical Image NetCDF toolkit, version 1.0.08 for Linux.^17^ DICOM files were converted to MINC-2 format and anonymized. All images were corrected for intensity non-uniformity using the N3 method with parameter optimization for 3T machines.^18,19^ MRI post-processing is illustrated in Figure 1. T2- or T1-weighted and FLAIR images were co-registered to the International Consortium for Brain Mapping (ICBM) 152 atlas using linear and non-linear techniques.^20,21^ For white matter segmentation, we used probabilistic tissue maps from the ICBM 152 atlas with a threshold of 95% probability, and only including supratentorial white matter.^22^ In order to avoid misclassification of juxtacortical white matter, we performed a further restriction of the white matter mask by applying consecutive operations of two erosions and one dilation,^23,24^ yielding a final mask that included mostly central white matter. White matter masks for each patient were obtained by inverse co-registration of atlas masks to the original individual images. Leukoaraiosis masks were obtained from FLAIR images combining manual outlining and signal thresholding. Leukoaraiosis signal threshold was defined as six standard deviations above the mean of splenium signal intensity, obtained from eight to ten 1 mm regions of interest. Normal-appearing white matter masks were then obtained by subtracting the leukoaraiosis mask from the white matter mask. For both leukoaraiosis and normal white matter segmentation, voxels below the maximum signal intensity in cerebrospinal fluid were excluded from the final masks, with this threshold obtained by sampling six to eight 1-mm regions of interest within the anterior horn of the lateral ventricles. In order to study the spatial relationship between leukoaraiosis burden and normal white matter signal, we also obtained normal white matter strata progressively distant from the leukoaraiosis borders by performing one to five recursive dilations of the leukoaraiosis mask, obtaining the difference of each one from the predecessor, and finally, intersecting the resulting mask with the normal white matter mask (*eg*., the white matter mask at a 3-voxel distance from leukoaraiosis was the result of the difference between the third leukoaraiosis dilation and the second leukoaraiosis dilation, intersected with the normal white matter mask). White matter signal intensity was normalized to splenium mean signal intensity obtained from the above mentioned splenium regions of interest.

**Figure 1.**
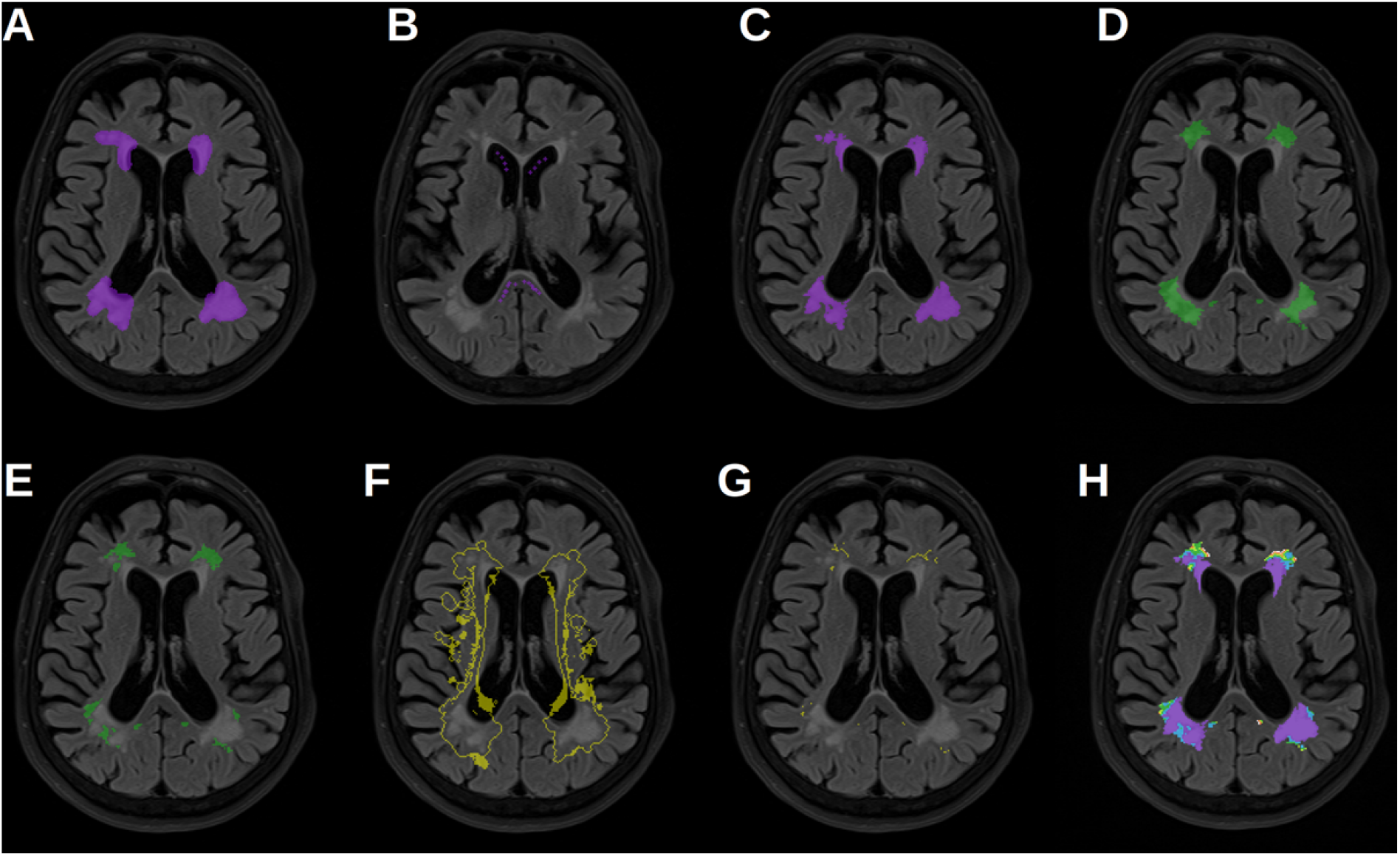
Segmentation of leukoaraiosis and normal appearing white matter. Legend. Leukoaraiosis segmentation was obtained by manually outlining a broad region of interest including all potential regions of leukoaraiosis (**A**). Secondly, eight to ten ROIs of 1 mm diameter were placed in the *splenium* and anterior horn of the lateral ventricles (**B**). The final leukoaraiosis mask (**C**) was obtained by thresholding the manually outlined mask to six standard deviations above the mean signal intensity obtained from the splenium regions. Subtracting the final leukoaraiosis volume from atlas-based white matter masks (**D**) resulted in the final normal appearing white matter mask (**E**). Voxels with values below the maximum value obtained from the lateral ventricles were excluded from the final masks of leukoaraiosis and normal-appearing white matter. The third one-voxel dilation of leukoaraiosis (**F**), which intersected with the final normal-appearing white matter mask (**E**) generated the normal-appearing white matter region at three-voxel distance from leukoaraiosis borders (**G**). Leukoaraiosis (purple) and all NAWM stratum according to voxel-distance from LKA (1, blue; 2, green; 3, yellow; 4, red; 5, white) are depicted (**H**).

Finally, the mean normalized FLAIR signal intensity of the normal appearing white matter (NAWM_M_) was obtained for each patient for statistical analysis, as well as the leukoaraiosis total volume (LKA_V_). For both leukoaraiosis and normal white matter segmentation, the infratentorial region was excluded. For patients with visible unilateral acute or chronic infarcts involving white matter, both NAWM_M_ and LKA_V_ were obtained from the contralateral hemisphere. In such cases, the total LKA_V_ measured in the unaffected hemisphere was doubled for statistical analysis.

### Statistical analysis

Data are reported as mean (± standard deviation), median (interquartile range), or frequency. Since most patients did not have sagittal imaging, LKA_V_ could not be normalized to intracranial volume, therefore we used LKA_V_ quartiles as an ordinal variable in the analysis. Univariate analysis of LKA_V_ and NAWM_M_ was performed using two-sample *t* or Wilcoxon test for categorical variables, and Pearson’s or Spearman’s correlation coefficient for quantitative variables, as appropriate. NAWM_M_ between 1-voxel dilation strata were compared with analysis of variance. The relationship between LKA_V_ and NAWM_M_ was studied by building linear and non-linear regression models using NAWM_M_ as the dependent variable and LKA_V_ as the independent variable. Multivariate regression analysis was performed including all variables associated with NAWM_M_ in univariate analysis, with stepwise backwards selection of final variables. This regression analysis was also performed separately for each of the 1-voxel dilation strata of normal white matter. Statistical significance was determined by *P*<0.05 on two-tailed tests. All statistical analyses was performed using R.

## Results

### Patients

Over the 36-month period, there were 317 cases of ischemic stroke or transient ischemic attack admitted to the hospital. Reasons for exclusion were absence of MRI (145), any combination of acute and/or chronic bilateral infarcts (43), absence of both T2- and T1-weighted imaging (24), and acquisition motion artifact (5), yielding a final sample of 100 patients (Table 1). LKA_V_ was positively associated with age (*ρ*=0.43; *P*<0.01), history of diabetes (*P*=0.03) and dementia (*P*<0.01).

**Table 1.** Study population.

|  | **Total, N=100** |
| --- | --- |
| Age | 68 ± 16 |
| Female sex | 53 |
| Hypertension | 68 |
| Diabetes | 32 |
| Body mass index | 27 ± 5 |
| Obesity | 19 |
| Atrial fibrillation | 7 |
| Heart failure | 2 |
| Ischemic heart disease | 13 |
| Prior statin use | 27 |
| Prior antiplatelet use | 29 |
| Prior anticoagulant use | 5 |
| Infarct location |  |
| Right hemisphere | 24 |
| Left hemisphere | 31 |
| Posterior fossa | 11 |
| No acute infarction | 34 |
| Leukaraiosis volume, cm<sup>3</sup> | 3 (2–16) |

Data are represented as mean ± standard deviation, frequency or median (interquartile range).

### NAWM_M_ analysis

NAWM_M_ was higher in the vicinity of leukoaraiosis borders and lower at stratum farther from leukoaraiosis (*P*<0.01 for analysis of variance; Figure 3). In univariate analysis, NAWM_M_ was associated with age (*r*=0.26; *P*=0.01), LKA_V_ (*ρ*=0.50; *P*<0.01) and history of heart failure (Table 2). In multivariable regression analysis, only LKA_V_ was independently associated with NAWM_M_ (Table 3).

**Table 2.** Group differences in mean FLAIR signal intensity on normal appearing white matter (NAWM_M_), univariate analysis.

|  | NAWM<sub>M</sub> |  | *P* |
| --- | --- | --- | --- |
|  | No | Yes |  |
| Female sex | 1.16 ± 0.07 | 1.14 ± 0.06 | 0.3 |
| Hypertension | 1.14 ± 0.06 | 1.15 ± 0.07 | 0.3 |
| Diabetes | 1.14 ± 0.06 | 1.16 ± 0.07 | 0.4 |
| Atrial fibrillation | 1.15 ± 0.06 | 1.17 ± 0.06 | 0.5 |
| Heart failure | 1.15 ± 0.06 | 1.24 ± 0.01 | <0.01 |
| Ischemic heart disease | 1.15 ± 0.05 | 1.16 ± 0.06 | 0.6 |
| Obesity | 1.15 ± 0.07 | 1.15 ± 0.05 | 0.9 |
| Dementia | 1.15 ± 0.06 | 1.16 ± 0.04 | 0.4 |
| Prior stroke | 1.15 ± 0.06 | 1.17 ± 0.07 | 0.3 |
| Prior statin use | 1.15 ± 0.06 | 1.14 ± 0.06 | 0.6 |
| Prior antiplatelet use | 1.15 ± 0.07 | 1.16 ± 0.06 | 0.4 |
| Prior anticoagulant use | 1.15 ± 0.06 | 1.12 ± 0.08 | 0.4 |

**Table 3.** Regression analysis of mean FLAIR signal intensity on normal appearing white matter (NAWM_M_).

|  | *B* | Confidence interval, 95% | *P* |
| --- | --- | --- | --- |
| **NAWM<sub>M</sub>, all voxels** |  |  |  |
| Leukoaraiosis quartile increase | 0.03 | 0.02–0.04 | <0.01 |
| Heart failure | 0.07 | -0.01—0.14 | 0.09 |
| **NAWM<sub>M</sub>, first voxel dilation**<br>**(voxels outlining leukoaraiosis)** |  |  |  |
| Leukoaraiosis quartile increase | 0.03 | 0.01–0.04 | <0.01 |
| **NAWM<sub>M</sub>, second voxel dilation** |  |  |  |
| Leukoaraiosis quartile increase | 0.24 | 0.11–0.37 | <0.01 |
| Leukoaraiosis quartile increase, second order term | -0.21 | -0.34–(-0.08) | <0.01 |
| **NAWM<sub>M</sub>, third voxel dilation** |  |  |  |
| Leukoaraiosis quartile increase | 0.22 | 0.10–0.34 | <0.01 |
| Leukoaraiosis quartile increase, second order term | -0.21 | -0.33–(-0.09) | <0.01 |
| **NAWM<sub>M</sub>, fourth voxel dilation** |  |  |  |
| Leukoaraiosis quartile increase | 0.19 | 0.07–0.31 | <0.01 |
| Leukoaraiosis quartile increase, second order term | -0.21 | -0.33–(-0.10) | <0.01 |
| **NAWM<sub>M</sub>, fifth voxel dilation** |  |  |  |
| Leukoaraiosis quartile increase | 0.19 | 0.07–0.30 | <0.01 |
| Leukoaraiosis quartile increase, second order term | -0.22 | -0.33–(-0.10) | <0.01 |

Voxel maps of leukoaraiosis frequency and NAWM_M_ are represented in Montreal Neurological Institute space in Figure 2. Patients with higher LKA_V_ had higher values of NAWM_M_, more conspicuously in the *centrum semiovale*. This increase in NAWM_M_ did not appear to correspond to frequency maps of leukoaraiosis and was also present in voxels not outlining leukoaraiosis regions. Indeed, in regression analysis, the association between NAWM_M_ and LKA_V_ was not restricted to voxels in the vicinity of leukoaraiosis. In voxels outlining leukoaraiosis (1-voxel dilation), NAWM_M_ showed a linear relationship with LKA_V_. In voxels not outlining leukoaraiosis (2-to 5-voxel dilations), there was a non-linear relationship with LKA_V_, with higher NAWM_M_ in patients in the second and third quartiles and lower values in the first and fourth quartiles of LKA_V_ (Figure 4; Table 3).

**Figure 2.**
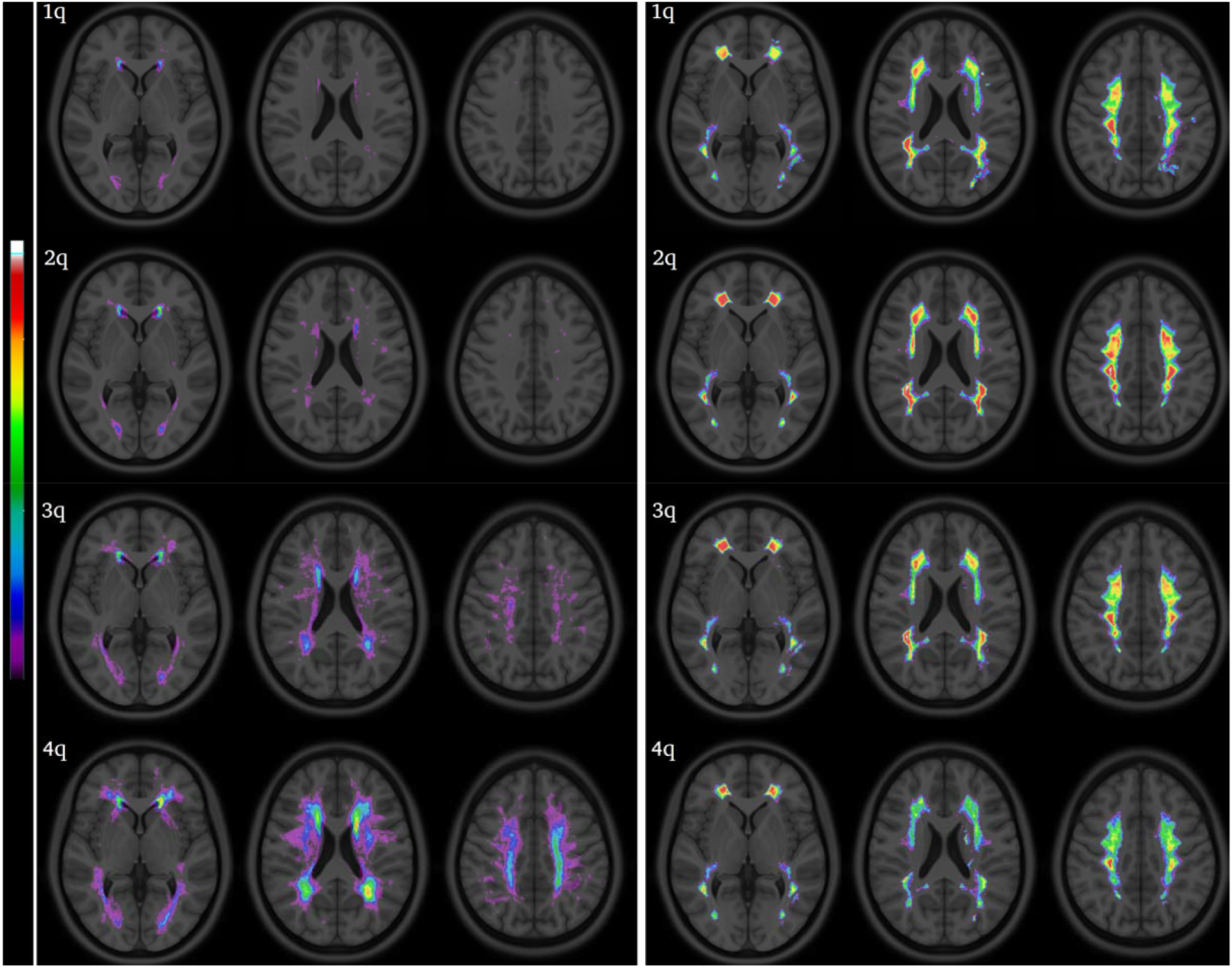
Voxel maps of leukoaraiosis and normal appearing white matter mean signal intensity on FLAIR in MNI space. Legend. Rows represent quartiles of leukoaraiosis volumes (1q—4q). The left panel represents leukoaraiosis frequency maps (spectral scale: 0–25). The right panel represents normal appearing white matter mean signal intensity in FLAIR (spectral scale: 0.2–1.2).

**Figure 3.**
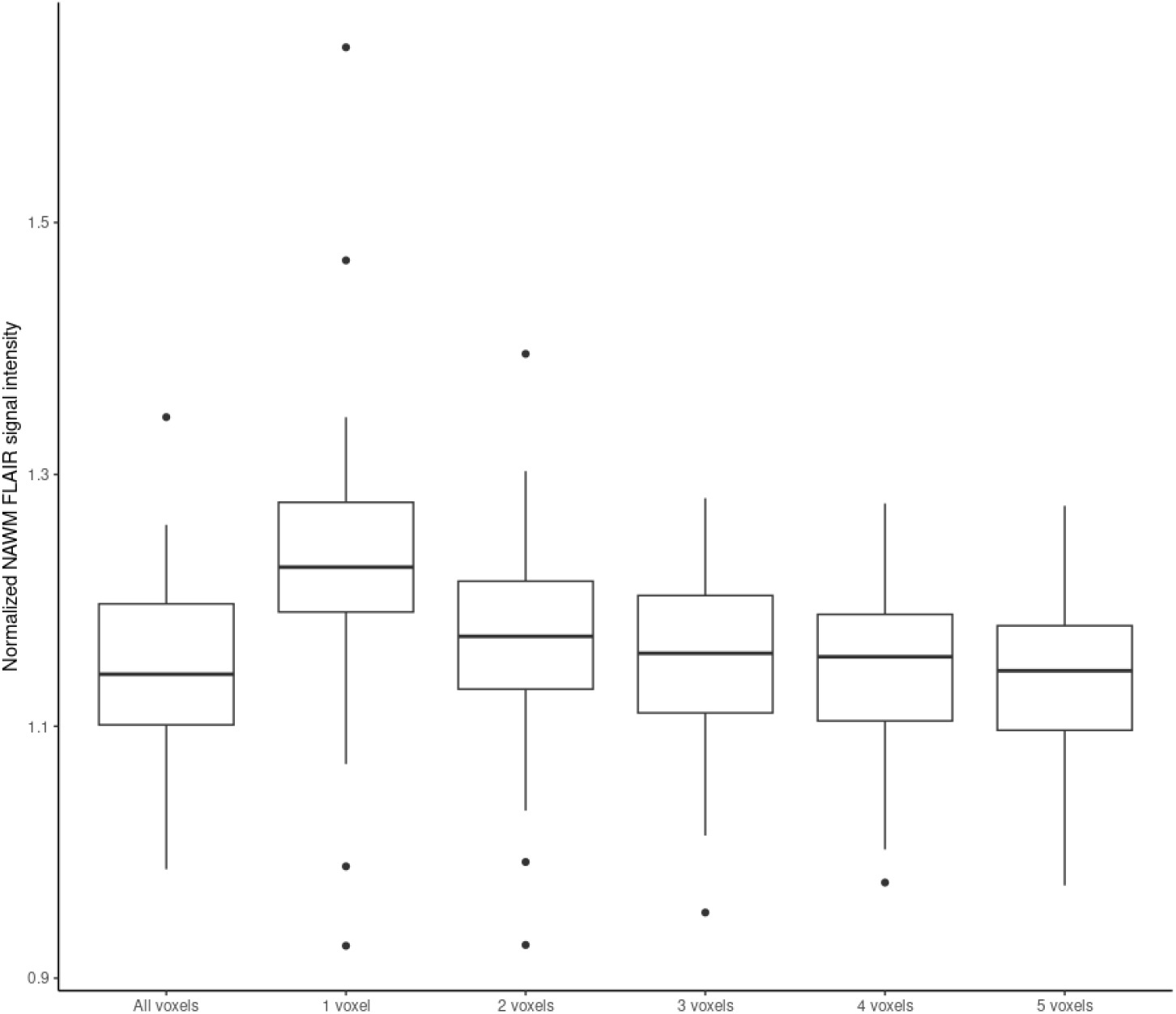
Normal appearing white matter mean signal intensity in FLAIR (NAWM_M_) according to voxel-distance from leukoaraiosis borders.

**Figure 4.**
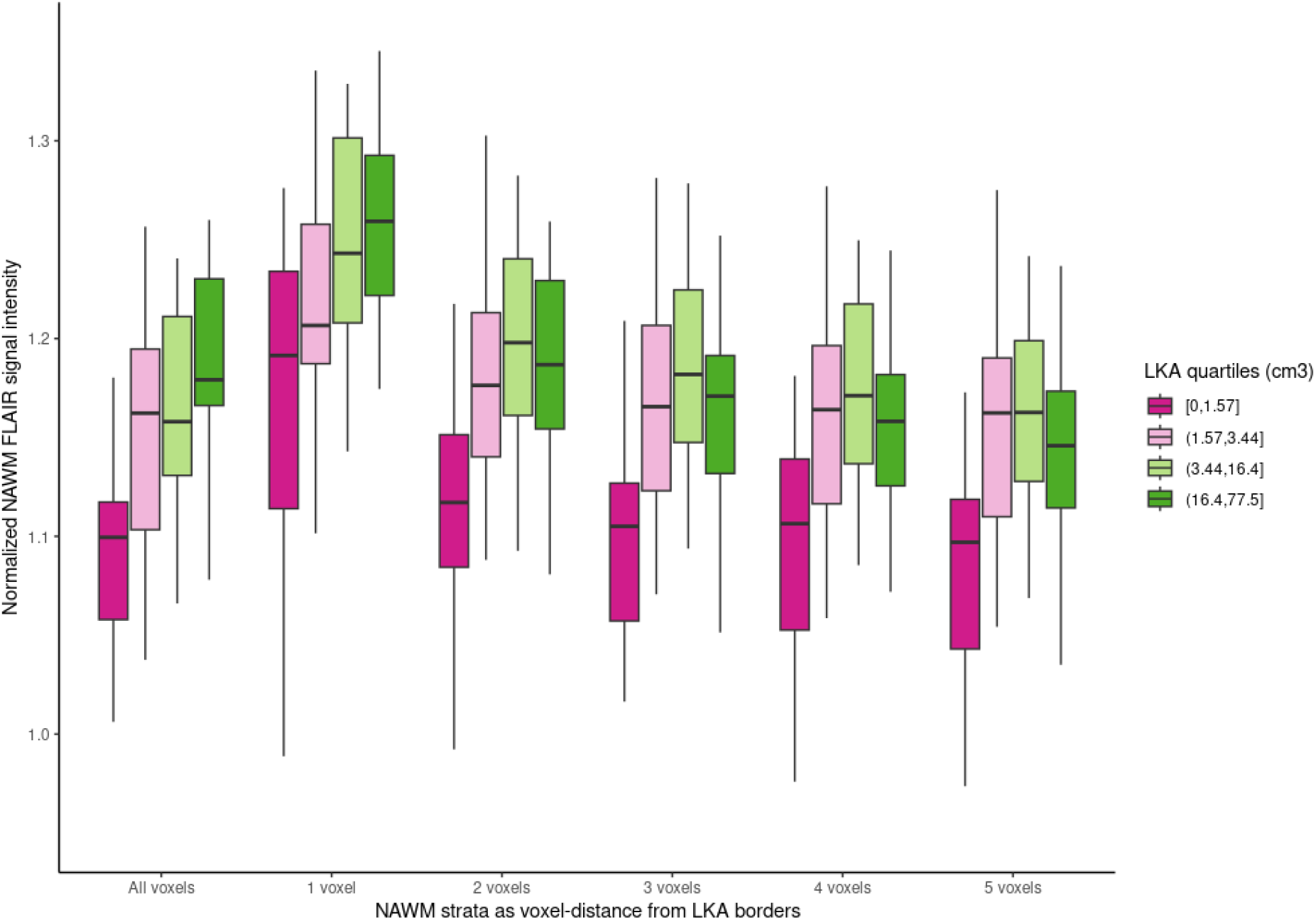
Normal appearing white matter mean signal intensity in FLAIR (NAWM_M_) at each leukoaraiosis (LKA) quartile, according to voxel-distance from leukoaraiosis borders.

## Discussion

In this study, we found that normal-appearing white matter presents subtle white matter intensity changes that are related to the burden of leukoaraiosis. The findings concur with the increasing evidence that white matter damage related to small-vessel disease is not restricted – and likely precedes – conspicuous leukoaraiosis. On the other hand, the topographical distribution of these signal intensity changes was not exclusively adjacent to those of leukoaraiosis, thus suggesting that it possibly reflect a diffuse process, and not only a milder, incipient form of leukoaraiosis.

Our results show a particular pattern of association between signal intensity in normal-appearing white matter and leukoaraiosis. First, signal changes related to leukoaraiosis were not restricted to the immediate borders (*ie*., 1-voxel dilation) of leukoaraiosis. Given that there are no formal criteria for defining the limits of leukoaraiosis, the presence of mild white matter hyperintensity at the borders of leukoaraiosis could be found as a result of a more conservative definition of leukoaraiosis related hyperintensity. However, we found that the association between leukoaraiosis volume and signal intensity in normal-appearing white matter was still present in white matter regions distant from leukoaraiosis limits. These results are in agreement with studies that demonstrate diffuse pre-visual changes in white matter in patients with small-vessel disease. Both pathology and DTI studies have shown that the typical signature of leukoaraiosis is also found in normal-appearing white matter.^11,13,14,25^ In pathology, this signature is characterized by demyelination, loosening of white matter fibers and accumulation of extracellular fluid.^26,27^ It has been proposed that the underlying etiology of leukoaraiosis stems from chronic endothelial damage leading to blood-brain barrier disruption and eventually to extravascular leakage and indiscriminate infiltration of brain tissue with blood constituents.^28,29^ We believe that such blood-brain barrier leakage could possibly account for the subtle FLAIR signal intensity changes observed on our study.

The topographical distribution of early white matter changes been controversial in the literature. The first study to address this issue described DTI changes to be in close proximity to large areas of leukoaraiosis, thus leading to the coining of the term “white matter hyperintensity penumbra”.^30^ DTI changes and baseline FLAIR intensity within a perimeter of 8mm distant from leukoaraiosis were predictors of incident leukoaraiosis in another study.^31^ On the other hand, Maniega *et al*. have demonstrated an association between leukoaraiosis severity and DTI markers of reduction of normal-appearing white matter integrity that was independent of proximity to leukoaraiosis.^25^ Also, studies evaluating markers of blood-brain barrier disruption have digressed from this proximity, penumbral pattern. In a longitudinal study with dynamic contrast-enhanced MRI in patients with Biswanger disease, blood-brain barrier disruption was related to baseline and incident leukoaraiosis, but the vast majority of voxels showing gadolinium enhancement were present not in leukoaraiosis regions, but within normal-appearing white matter, and not in close proximity of either baseline or newly formed leukoaraiosis regions.^32^ Therefore, it appears that leukoaraiosis related changes might develop diffusely across brain white matter, and not exclusively in a penumbral pattern around established leukoaraiosis. Our results are in agreement with that hypothesis. We believe that subtle signal intensity changes on FLAIR, as well as DTI and blood-brain barrier changes, might not represent a milder stage of leukoaraiosis, but rather reflect widespread etiopathogenic processes underlying small-vessel disease and thus, leukoaraiosis development.

Interestingly, we found that the pattern of association between leukoaraiosis volume and FLAIR signal intensity was different between 1-dilation masks and 2-to 5-dilation masks. In voxels not outlining the leukaraiosis region (*i*.*e*., within the 2-to 5-dilation masks), patients in the highest quartile burden of leukoaraiosis showed a decrease when compared to patients in the third quartile, yielding a non-linear association between NAWM_M_ values and leukoaraiosis volume. We hypothesize that, if subtle signal intensity increase represents early etiopathogenic processes underlying leukoaraiosis development, this decrease of signal intensity in patient with very high volumes of leukoaraiosis might reflect end-stage or “burnout” white matter damage. Alternatively, further signal intensity increase in the fourth quartile was absent possibly because 2-to 5-dilation masks would be closer to juxtacortical regions among these patients with very high leukoaraiosis volumes, where leukoaraiosis related changes might be less pronounced or absent.

Despite being related to leukoaraiosis, FLAIR signal intensity in normal-appearing white matter was not related to the established risk factors for leukoaraiosis, such as age and hypertension. FLAIR signal intensity in normal-appearing white matter was related to history of cardiac failure in univariate analysis, however there were only two patients with this condition in our sample. We hypothesize that in patients with heart failure, increase in FLAIR signal intensity could be a result of increased capillary pressure and mild extravascular leakage. If, as mentioned before, leukoaraiosis and related changes reflect underlying blood-brain barrier disruption, this effect of heart failure on FLAIR signal intensity could be enhanced in patients with more severe leukoaraiosis, however an interaction analysis could not be performed in our sample due to the small sample of patients with history of heart failure.

Our study has some limitations. Our data derives from MRI acquired for standard clinical care, with suboptimal resolution and absence of DTI, perfusion imaging or gadolinium-enhanced sequences. Thus, the association of our findings with other putative markers of white matter damage could not be assessed. Also, we were not able to correct leukoaraiosis for intracranial volume with the available MRI data, and analysis was thus performed using volume quartiles. In addition, to minimize tissue misclassification, we selected a conservative white matter mask for segmentation, which excluded juxtacortical white matter. Finally, as a retrospectively collected sample, we could not study the relationship of our findings with stroke outcomes.

In conclusion, the burden of leukoaraiosis is associated with subtle, diffuse FLAIR signal intensity changes in normal-appearing white matter. Further studies are necessary to assess neuroimaging and pathology correlates and the clinical significance of these changes.

## Data Availability

The data that support the findings of this study are available on reasonable request to the corresponding author.

## Non-standard Abbreviations and Acronyms

ICBM: International Consortium for Brain Mapping.
DTI: diffusion tensor imaging.
FLAIR: fluid-attenuated inversion recovery.
LKA_V_: leukoaraiosis volume.
NAWM_M_: mean FLAIR signal intensity in normal-appearing white matter.

## Funding

Octavio Pontes-Neto has received support from the National Council for Scientific and Technological Development (CNPq) grant #311916/2022-8.

## Disclosures

The authors report no competing interests.

## Notes

### Competing Interest Statement

The authors have declared no competing interest.

### Author Declarations

This project was approved by the Research Ethics Committee of the Hospital Pró-Cardíaco, Rio de Janeiro, Brazil.

